# Serum bile acids improve prediction of Alzheimer’s progression in a sex-dependent manner

**DOI:** 10.1101/2022.12.26.22283955

**Authors:** Tianlu Chen, Lu Wang, Guoxiang Xie, Xiaojiao Zheng, Bruce S. Cristal, Tao Sun, Matthias Arnold, Mengci Li, Siamac Mahmoudian Dehkordi, Matthew J. Sniatynski, Qihao Guo, Lirong Wu, Junliang Kuang, Jieyi Wang, Kwangsik Nho, Zhenxing Ren, Alexandra Kueider-Paisley, Rima Kaddurah-Daouk, Wei Jia, the Alzheimer’s Disease Neuroimaging Initiative, the Alzheimer Disease Metabolomics Consortium

## Abstract

**INTRODUCTION:** There is evidence that there are differences in the serum levels of bile acids (BAs) in males and females and their risk of developing Alzheimer’s disease (AD). We previously reported that serum BAs are associated with AD. It remains unclear, however, how changes in serum BAs may relate to the development of AD in a sex-dependent manner.

**METHODS:** We analyzed 33 BAs in the sera of 4219 samples from 1180 subjects in the ADNI cohort. Using linear models, we examined the associations between BAs and mild cognitive impairment (MCI) progression and clinical markers.

**RESULTS:** Significant alterations in BA profiles occurred at an early stage of MCI and were associated with the onset and progression of MCI. These changes were more dramatic in men than in women. BA markers improved the ability of current clinical markers to diagnose MCI and predict its progression.

**DISCUSSION:** Our results highlight the role of BAs in the development of AD and may help improve AD prediction and personalized therapies.

**Research in context:**

1. **Systematic review:** We examined the relationship between bile acid (BA), mild cognitive impairment (MCI), and Alzheimer’s disease (AD). We previously reported this association. Our findings were consistent with those of other studies, although previous research did not consider sex differences or comprehensively evaluate the potential of BAs as diagnostic markers for AD.
2. **Interpretation**: Our results suggest that changes in BA profiles may play a role in the development of AD and that sex-specific differences may be important for personalized prediction and management of the disease.
3. **Future directions**: In the future, it will be important to confirm our findings with other independent samples and further investigate the ways in which BA metabolism, including cholesterol catabolism in the liver and brain, may contribute to AD.

## 1. Introduction^1^

Alzheimer’s disease (AD) affects millions of people and puts a growing strain on healthcare systems around the world. To date, there have been no effective therapies for preventing or slowing the progression of AD. Epidemiological studies have confirmed differences in the risk and severity of AD between men and women, but the underlying mechanisms are only beginning to be understood [1, 2]. Metabolomics, the study of small molecules in living organisms, has the potential to identify biomarkers and understand the development of mild cognitive impairment (MCI) and AD [3-6]. In our previous studies, we used targeted [7] and untargeted [4] metabolomics to identify metabolic signatures related to disease and disease progression in serum and brain samples from the Alzheimer’s Disease Neuroimaging Initiative (ADNI) and Religious Orders Study and Rush Memory and Aging (ROS-MAP) cohorts. We also examined the effect of sex on the relationship between metabolic changes and AD, which may help explain the observed differences in AD susceptibility and severity between men and women [8].

Bile acids (BAs), a group of metabolites involved in cholesterol catabolism, have received significant attention in recent years due to their important biological characteristics and functions [9, 10]. Primary BAs (CA and CDCA) are synthesized in hepatocytes from cholesterol, while secondary BAs are biotransformed by gut bacteria from primary BAs. As a result, changes in BA profiles may reflect changes in cholesterol metabolism and bacterial functions, both of which have been linked to the development and progression of AD. Previous research has shown that BA profiles are significantly altered in patients with mild cognitive impairment (MCI) and AD compared to individuals with normal cognition (NC)[11-14]. Higher levels of secondary BAs and their conjugated forms and higher ratios of secondary to primary BAs are associated with worse cognitive function and amyloid-beta, tau, and neurodegenerative (A/T/N) biomarkers [12, 13]. Lower levels of primary BAs have been linked to increased brain amyloid deposits, faster accumulation of white matter lesions, and increased brain atrophy [15]. We have previously suggested that changes in BA profiles may play a role in the development of AD and that the gut microbiome-bile acid-brain cholesterol axis may be a potential target for the prevention and treatment of AD [14, 16]. In addition, we and others have observed that BA profiles are affected by sex, and sex differences should therefore be considered when studying BA changes related to diseases [17, 18].

In this study, we examined the concentration of BAs in multiple independent samples. In addition to the concentration of BAs, we also analyzed the percentages of individual BAs relative to the total concentration of BAs (TBA) and calculated ratios that reflect the enzymatic activities and functions of the gut microbiota. We identified the most representative BA features that demonstrated the best overall performance and analyzed their performance and alteration over time in relation to sex and the development of AD. We also comprehensively evaluated the potential of these BA features to improve diagnostic and predictive markers for AD. Our goal is to replicate and refine previous findings, test hypotheses, and increase our understanding of the gut microbiome-bile acid-brain cholesterol axis and its role in AD.

## 2. Methods

### 2.1. Study cohorts and sample collection

Baseline and follow-up serum samples of 1180 individuals and related information were obtained from the ADNI study. Written informed consent was obtained at the time of enrollment which was approved by each participating sites’ institutional review board. Complete information on ADNI study including inclusion and exclusion criteria, sample collection protocol, and clinical marker extraction (demographics, apolipoprotein E ε4 genotype (APOE-4), neuropsychological test scores, cerebrospinal fluid (CSF) and positron emission tomography (PET) imaging markers) can be found at http://adni.loni.usc.edu/data-samples/access-data/.

### 2.2. Quantitative measurement and pretreatment of metabolic data

Targeted metabolites (nN104), including 33 BAs, 41 free fatty acids (FA), and 30 amino acids and organic acids, were quantitatively measured and processed using an ultra-performance liquid chromatography coupled to tandem mass spectrometry system (UPLC-MS/MS) [19] and the TMBQ software (V1.0, HMI, Shenzhen, China). Sixty-nine extended BA features (Table S1) were generated comprising concentration percentages of each BA to TBA and BA ratios reflective of enzymatic activities and gut microbiota functions. All the metabolic variables were logarithmic transformed for statistical analysis.

### 2.3. Clinical marker panels

Six types of clinical markers were included in the subsequent analysis, namely, a cognition score panel with 22 neuropsychological test scores, a CSF panel with five A/T markers derived from CSF, an AV45-PET panel comprising three PET imaging markers indicating brain amyloid-beta deposition, an FDG-PET panel comprising five 18F-fluorodeoxyglucose PET imaging markers associated with brain D-glucose metabolism, a demographic panel comprising age, BMI, and education year, and an APOE-4 genotype status marker.

### 2.4. Statistical analyses

To obtain higher statistical power, baseline and follow-up samples were pooled (total nN4219). Differences in clinical markers were evaluated using Chi-squared test, student’s t-test, Mann-Whitney test, Analysis of variance, or Kruskal–Wallis test followed by Dunn’s multiple-comparison post-hoc, as appropriate and as denoted in the text and figure legends. Principle component analysis (PCA) and locally weighted regression (LOESS) were used for dimension reduction and curve fitting of marker panels. Mixed linear models were fitted to examine the association of BA features with diagnostic groups (normal cognition (NC), early mild cognitive impairment (EMCI), late mild cognitive impairment (LMCI), and AD) after adjusting fixed and random effects. Logistic regression was used to identify features associated with disease progression from MCI to AD after adjusting confounders. Partial Pearson’s correlation was used for association analysis between BA features and clinical markers after adjusting confounders. Two linear regression models were used to select medications for adjustment for each BA feature (Table S2). The contribution of BA features to clinical practice were assessed according to the improvement of AUC derived from logistic regression models based on each type of clinical marker panel alone or on clinical marker panel combined with four selected BA features.

All the data analyses were conducted using R (V3.5.1) and GraphPad (V9.3). All p-values were adjusted using the Benjamini–Hochberg’s false discovery rate (FDR) and the significance level was 0.05 (two-tailed) unless otherwise indicated. Further details are provided in the supplementary information.

## 3. Results

### 3.1. Study samples

A total of 4219 pooled serum samples (2386 from men and 1833 from women; median (min, max) age of 75 (55, 95) years old) were involved in this study. The samples were drawn from 1180 individuals at baseline and throughout their 6–120 months follow-up (mean follow-up duration was 33.2 months) (Table 1 and S3 and Figure 1a). In total, 35.8%, 27.4%, and 47.8% samples have clinical markers of CSF, AV45-PET, and FDG-PET tests. Among them, 1259, 691, 1130, and 1139 samples were diagnosed with NC, EMCI, LMCI, and AD respectively. Of the 1821 MCI samples, 537 were later converted to AD (cMCI), whereas 1284 were sustained MCI (sMCI; Table S4).

**Table 1.**
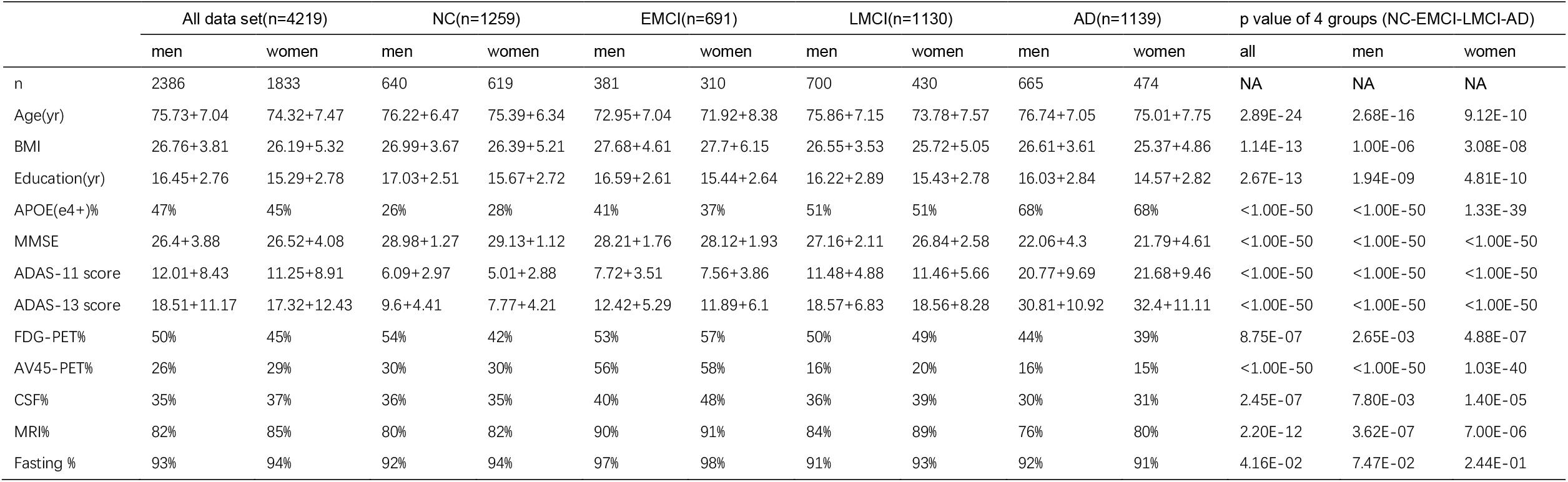
Demographic and clinical characteristics of the 4219 samples included in this study.

|  | All data set(n=4219) |  | NC(n=1259) |  | EMCI(n=691) |  | LMCI(n=1130) |  | AD(n=1139) |  | p value of 4 groups (NC-EMCI-LMCI-AD) |  |  |
| --- | --- | --- | --- | --- | --- | --- | --- | --- | --- | --- | --- | --- | --- |
|  | men | women | men | women | men | women | men | women | men | women | all | men | women |
| n | 2386 | 1833 | 640 | 619 | 381 | 310 | 700 | 430 | 665 | 474 | NA | NA | NA |
| Age(yr) | 75.73+7.04 | 74.32+7.47 | 76.22+6.47 | 75.39+6.34 | 72.95+7.04 | 71.92+8.38 | 75.86+7.15 | 73.78+7.57 | 76.74+7.05 | 75.01+7.75 | 2.89E-24 | 2.68E-16 | 9.12E-10 |
| BMI | 26.76+3.81 | 26.19+5.32 | 26.99+3.67 | 26.39+5.21 | 27.68+4.61 | 27.7+6.15 | 26.55+3.53 | 25.72+5.05 | 26.61+3.61 | 25.37+4.86 | 1.14E-13 | 1.00E-06 | 3.08E-08 |
| Education(yr) | 16.45+2.76 | 15.29+2.78 | 17.03+2.51 | 15.67+2.72 | 16.59+2.61 | 15.44+2.64 | 16.22+2.89 | 15.43+2.78 | 16.03+2.84 | 14.57+2.82 | 2.67E-13 | 1.94E-09 | 4.81E-10 |
| APOE(ε4+)% | 47% | 45% | 26% | 28% | 41% | 37% | 51% | 51% | 68% | 68% | <1.00E-50 | <1.00E-50 | 1.33E-39 |
| MMSE | 26.4+3.88 | 26.52+4.08 | 28.98+1.27 | 29.13+1.12 | 28.21+1.76 | 28.12+1.93 | 27.16+2.11 | 26.84+2.58 | 22.06+4.3 | 21.79+4.61 | <1.00E-50 | <1.00E-50 | <1.00E-50 |
| ADAS-11 score | 12.01+8.43 | 11.25+8.91 | 6.09+2.97 | 5.01+2.88 | 7.72+3.51 | 7.56+3.86 | 11.48+4.88 | 11.46+5.66 | 20.77+9.69 | 21.68+9.46 | <1.00E-50 | <1.00E-50 | <1.00E-50 |
| ADAS-13 score | 18.51+11.17 | 17.32+12.43 | 9.6+4.41 | 7.77+4.21 | 12.42+5.29 | 11.89+6.1 | 18.57+6.83 | 18.56+8.28 | 30.81+10.92 | 32.4+11.11 | <1.00E-50 | <1.00E-50 | <1.00E-50 |
| FDG-PET% | 50% | 45% | 54% | 42% | 53% | 57% | 50% | 49% | 44% | 39% | 8.75E-07 | 2.65E-03 | 4.88E-07 |
| AV45-PET% | 26% | 29% | 30% | 30% | 56% | 58% | 16% | 20% | 16% | 15% | <1.00E-50 | <1.00E-50 | 1.03E-40 |
| CSF% | 35% | 37% | 36% | 35% | 40% | 48% | 36% | 39% | 30% | 31% | 2.45E-07 | 7.80E-03 | 1.40E-05 |
| MRI% | 82% | 85% | 80% | 82% | 90% | 91% | 84% | 89% | 76% | 80% | 2.20E-12 | 3.62E-07 | 7.00E-06 |
| Fasting % | 93% | 94% | 92% | 94% | 97% | 98% | 91% | 93% | 92% | 91% | 4.16E-02 | 7.47E-02 | 2.44E-01 |

**Figure 1.**
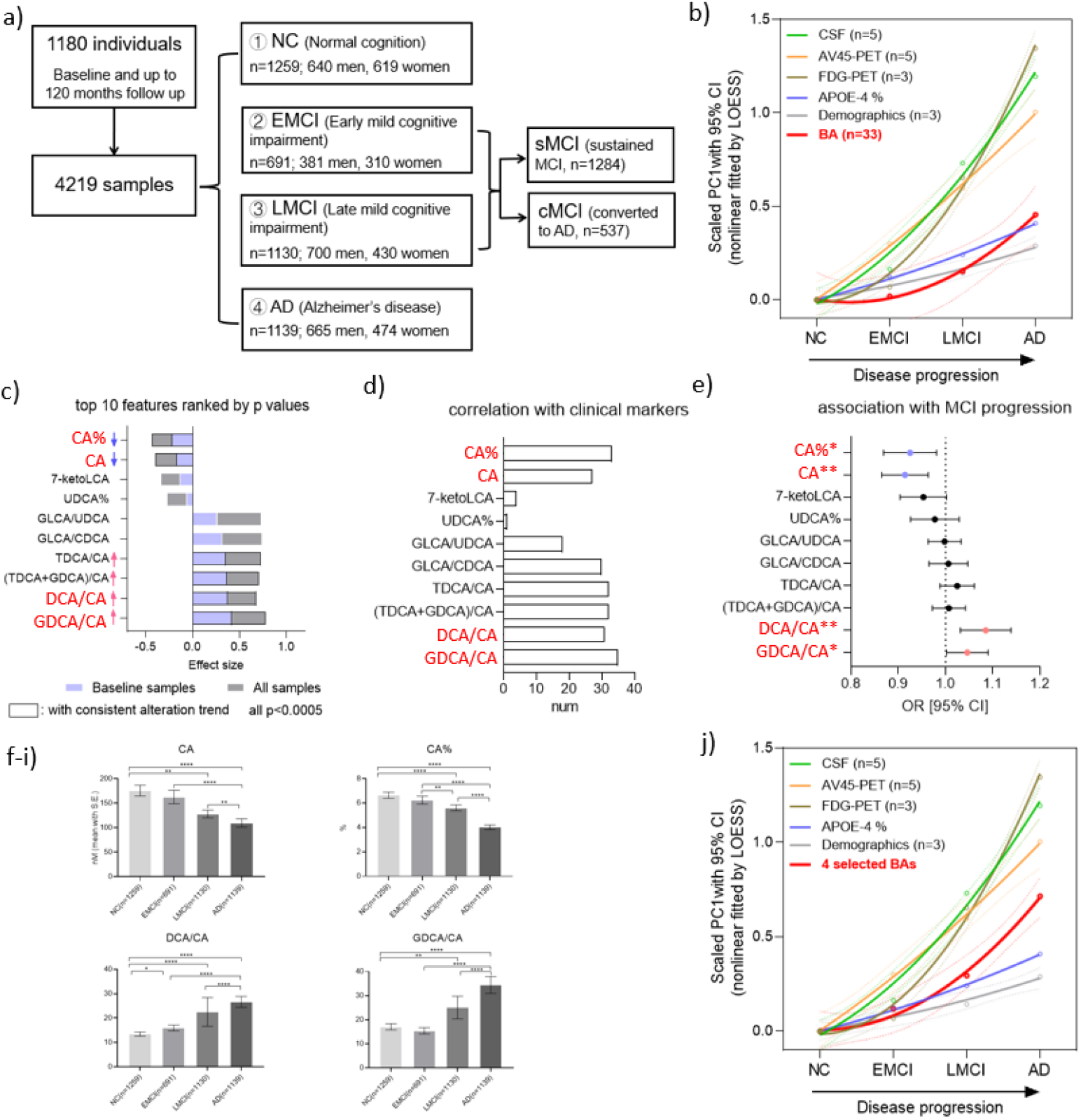
a) Study samples stratified by diagnostic groups and disease progression. b) Dynamic changes (LOESS fitted curves with 95% CI of PC1, the first component of PCA) of clinical marker panels and BAs in diagnostic groups (NC, EMCI, LMCI, and AD). Changes in BAs are observed early in the development of AD and continue across the spectrum of disease severity, which is comparable to those of APOE-4 genotype proportion and demographics. c) Bar plot of effect sizes (coefficients) of 10 BA features derived from separate linear mixed models, with diagnostic group as dependent variable and each of the z-scored BA features as independent variables respectively. Top 10 BA features with lowest *P*-values based on baseline and all samples are listed and are ordered by their effect size based on baseline samples. The alteration trend is labeled if it continues to rise or fall, in four diagnostic groups. d) Number of correlations (FDR<0.05) between the 10 BA features and 35 clinical markers. e) *P*-values and OR (95% CI) for 10 BA features from logistic regression for the prediction of MCI progression. *, FDR < 0.05; **, FDR < 0.01. f-i) Levels (mean with S.E.) of CA, CA%, DCA/CA, and GDCA/CA in NC, EMCI, LMCI, and AD groups. j) Dynamic changes of clinical marker panels and four selected BA features with disease progression.

### 3.2 Serum BA profiles were associated with AD development

PCA was conducted on all serum metabolites and the first component, PC1, was taken as the representative variable. Changes of serum metabolites (PC1) were observed early in AD development and continued across the spectrum of disease severity (Figure S1). We then evaluated alterations of BAs, FAs, and other metabolites in the same way. BAs increased continuously suggesting its association with AD progression (Figure S1). Compared with clinical marker panels, the alterations of BAs were comparable to those of APOE-4 proportion and demographics, which were less apparent than those of CSF and PET panels, throughout four diagnostic groups (Figure 1b). These findings demonstrated that alterations in serum metabolites, particularly BAs, were associated with diagnostic groups of AD.

### 3.3. Four BA features were identified as potential AD markers

More BA features were generated based on the 33 BAs (Table S1). Four were identified as potential AD markers based on their alteration trends and associations with diagnostic groups, clinical markers, and disease progression. Ten features that differed most significantly (with the lowest P-values derived from Mixed Linear Models) among NC, EMCI, LMCI, and AD in pooled and baseline samples are listed in Figure 1c. Of these, CA and CA% levels decreased, whereas TDCA/CA, (TDCA+GDCA)/CA, DCA/CA and GDCA/CA levels increased continuously across four groups. These BA features showed associations with more clinical markers than others (Figure 1d). Logistic regression analysis revealed that four of them were association with disease progression from MCI to AD (Figure 1e). None of them were associated with the progression from NC to MCI. Accordingly, four BA features, CA, CA%, DCA/CA, and GDCA/CA, were considered representative of BA profiles (Figure 1f-1i). As expected, changes of these four features were more apparent than those of APOE-4 proportion and demographics across four diagnostic groups (Figure 1j).

### 3.4 BA features were more sensitive in men than in women

Stratification analysis on sex was performed on the four selected features because BA profiles are known to be affected by sex. Consistent to previous findings that TBA was higher in men than in women (Figure S2). Earlier and more apparent alteration trends and larger differences between groups were observed in men than in women (figure 2a-2e). Correlation analysis of the four BA features and clinical markers showed that more correlated pairs were detected in men than in women (figures 2f and 2g). During the 120-month follow-up period, 336 male and 201 female samples converted from MCI to AD and 745 male and 539 female samples sustained in the MCI stage. Logistic regression analysis indicated that these features were associated with progression from MCI to AD in men but not in women (table S5). These results signify that the changes of four BA features were more dramatic in men than in women.

**Figure 2.**
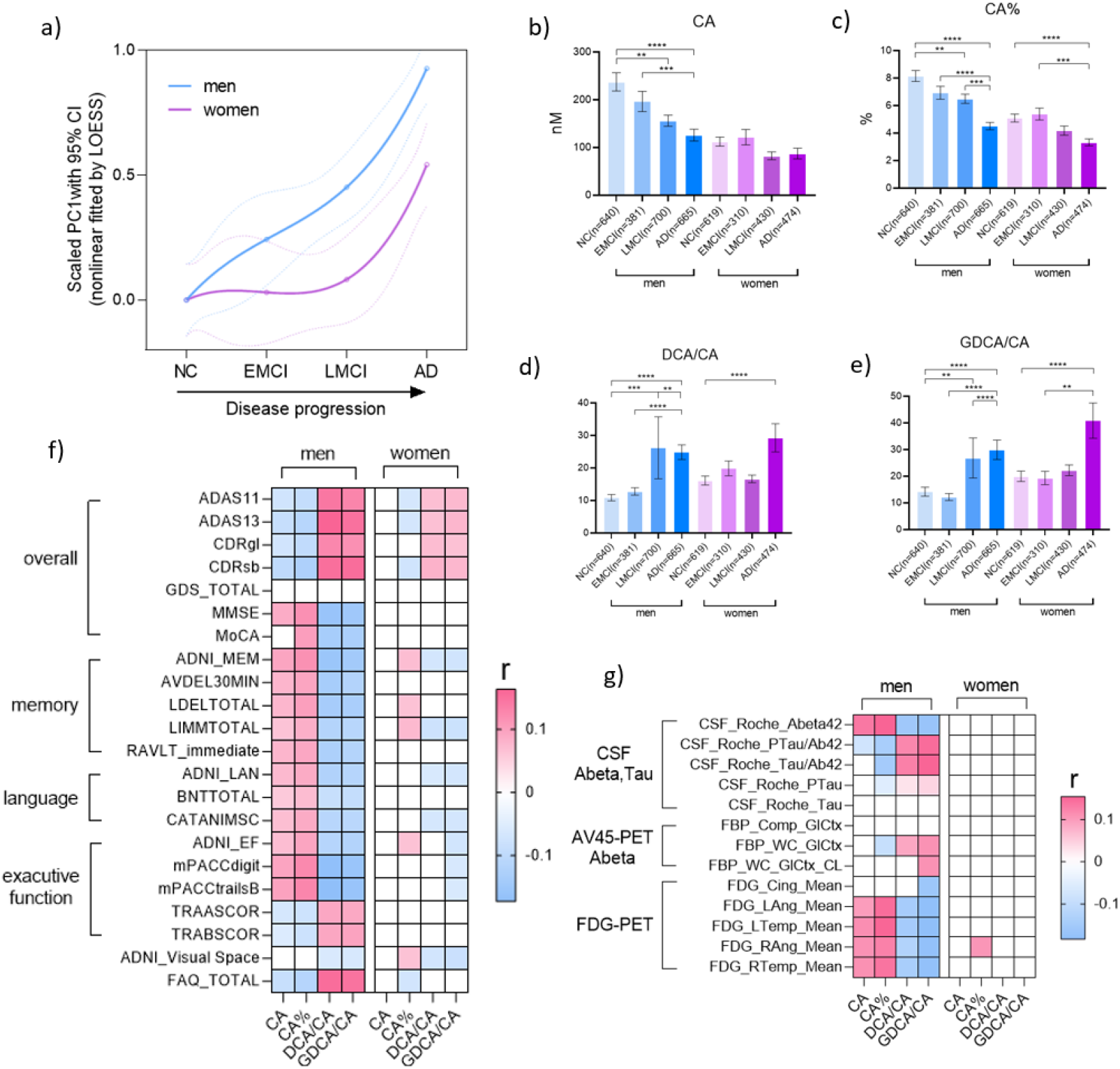
Dynamic changes of four selected BA features with disease progression in men and women. Changes in men are earlier and more apparent than that of women. b-e) Levels (mean with S.E.) of the four BA features in NC, EMCI, LMCI, and AD groups in men and women. *P*-values correspond to comparisons using the Kruskal–Wallis test with Dunn’s post-hoc comparisons. Heatmaps of the correlation coefficients between the four features and clinical markers from neuropsychological tests (e) and CSF and PET imaging (f) in men and women. Blank, non-significant (FDR>0.05); pink, positive association; blue, negative association.

### 3.5 BA features were associated with MCI progression speed in men

During the 120-month follow-up period, 103 male samples (from 53 subjects) converted from NC to EMCI or LMCI, 336 male samples (from 176 subjects) converted from EMCI or LMCI to AD, and 349 male samples (from 179 subjects) were newly diagnosed with AD (336 progressed from MCI and 13 progressed from NC) (Table S6). According to conversion time, these samples were stratified into 15 time groups (Figure 3a) and levels of the four BA features at these groups were plotted (Figures 3b–e). Interestingly, for MCIs (the yellow panel), the levels of CA and CA% decreased rapidly until 48 months before progression to AD and were then maintained at a low level. DCA/CA and GDCA/CA levels increased roughly with the speed of MCI progression. As no comparisons of the four features between any two time groups reached statistical significance, we (post-hoc) combined the seven time groups into two time periods (≤48 months and >48 months) to increase statistical power. Levels of the four features were significantly different between the two periods and also between the MCIs who progressed to AD in ≤48 months and sMCIs (Figures 3f-3i). Taken together, the four BA features were associated with the speed of MCI progression in men and have potential to identify those MCI patients likely to progress to AD in 4 years.

**Figure 3.**
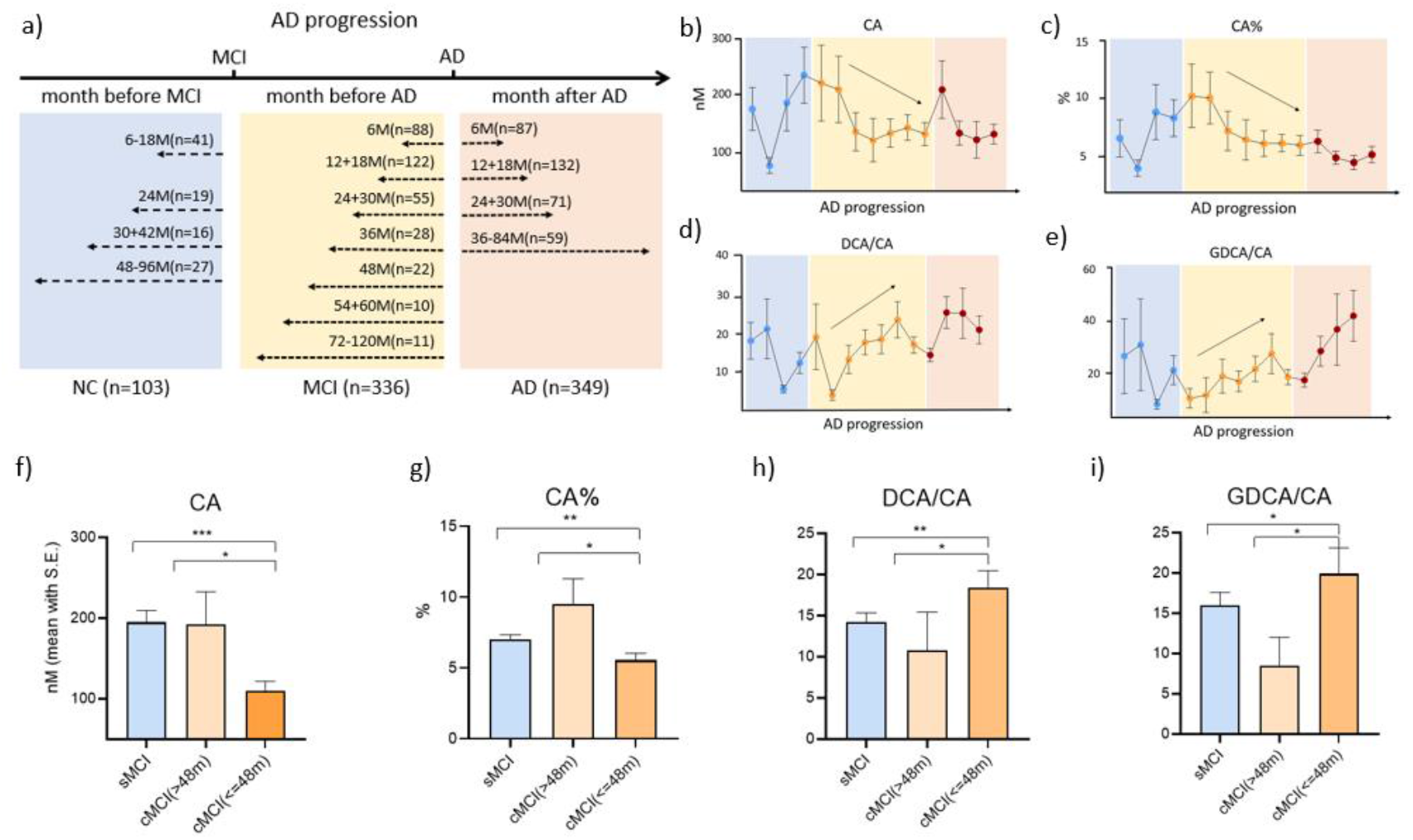
a) Fifteen time groups of AD progression trajectory, including four time groups for NCs before they progressed to MCI (blue panel), seven time groups for samples from subjects with MCI before they progressed to AD (yellow panel), and four time groups for samples from subjects with AD after they were diagnosed with AD (pink panel). Trajectories (mean with S.E.) of CA (b), CA% (c), DCA/CA (d), and GDCA/CA (e) across AD progression. Levels (mean with S.E.) of CA (f), CA% (g), DCA/CA (h), and GDCA/CA (i) in sMCI, and in cMCI patients who progressed to AD in >48 or in <N48 months. *, significant difference based on the Kruskal– Wallis test with Dunn’s post-hoc comparisons.

### 3.6 BA features improved the performances of clinical markers for prediction of MCI progression in men

Motivated by above findings, we assessed the contributions of the four BA features to clinical marker panels for diagnosis and progression prediction in men. The AUCs of diagnostic models (logistic regression) based on each type of clinical marker panels and on their combination with four BA features to discriminate between NC-MCI, NC-AD, and MCI-AD are listed in Table S7. It was observed that 13 of the 15 models had increased AUCs after the addition of BA features indicating the positive contributions of BA features to existing clinical marker panels. The cumulative AUC improvements to APOE-4 (21.6%) and demographics (17.6%) were higher than other marker panels (Figure 4a). The addition of BA features also led to positive contributions to clinical marker panels (improved AUCs ranging 1.1%-7.0%) for the discrimination between sMCI and cMCI patients (Figure 4b and Table S8). Comparatively, larger improvements were also observed in APOE-4 (7.0%) and demographics (5.3%). Furthermore, we examined the contributions of BA features to existing clinical marker panels, for the discrimination between sMCI and cMCI samples who converted to AD in different durations (in 0-1, 0-2, 0-3, and 0-4 years). Greater improvements were observed in samples acquired closer to the time of conversion, for APOE-4 and demographics (Table S8).

**Figure 4.**
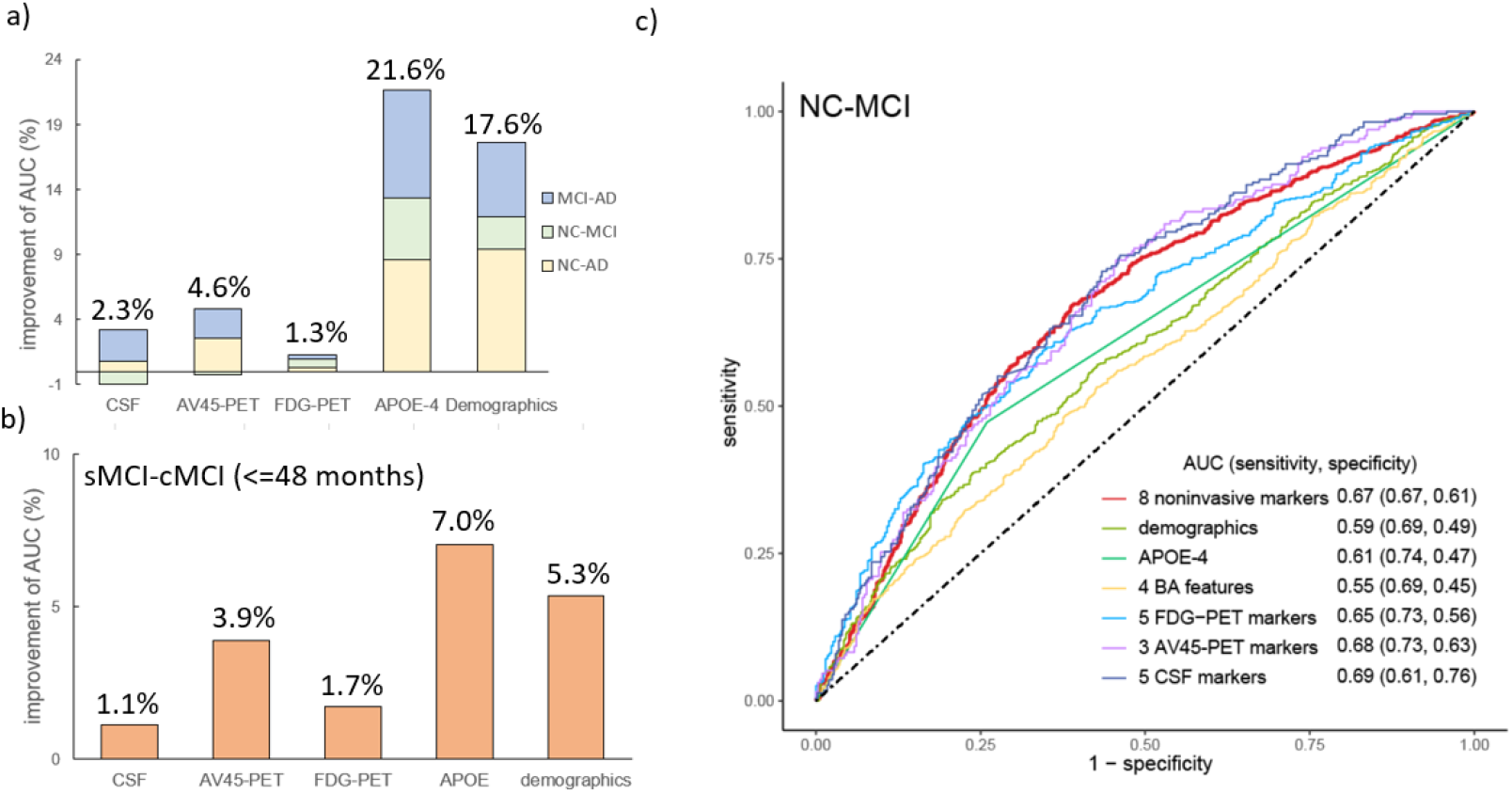
Improvements in the AUC values (%) of logistic regression models using each type of clinical markers following the addition of BA features for discriminating NC-AD, NC-MCI, and MCI-AD (a) and for discriminating sMCI and cMCI patients who progressed to AD within 4 years (b). c) ROC curves for the NC-MCI diagnostic models using eight noninvasive markers (age, BMI, education year, APOE-4, CA, CA%, DCA/CA, and GDCA/CA), four BA features, and different types of clinical markers.

Given the contributions of BA features to APOE-4 and demographics, a series of diagnostic and predictive models (logistic regression) were constructed using eight non-invasive markers (age, BMI, education year, APOE-4, CA, CA%, DCA/CA, and GDCA/CA) in men (Table S9). Notably, the performance of our model in discriminating NC and MCI (AUC, 0.67; sensitivity, 0.67; specificity, 0.61; red curve) was comparable to that of models based on CSF (AUC, 0.69; sensitivity, 0.61; specificity, 0.76), AV45-PET (AUC, 0.67; sensitivity, 0.63; specificity, 0.73), and FDG-PET (AUC, 0.65; sensitivity, 0.56; specificity, 0.73) markers (Figure 4c). The AUCs of the predictive models for discriminating sMCI and cMCI were higher in samples from patients who converted to AD at a faster pace (Table S9).

## 4. Discussion

There is growing evidence that neurodegenerative diseases, such as AD, are closely linked to changes in the serum metabolome. Our study is one of the first to examine the relationship between BAs and AD development [12, 13, 15]. Our findings highlight the sexually dimorphic roles of BAs in AD development and suggest their potential for clinical use.

It is well-recognized that different types of markers have different alteration patterns and paces in the evolution of AD [20]. This study characterized trajectories of 104 serum metabolites, 33 BAs, and four selected BA features during AD development. Our data indicated that the changes in BAs (Figure 1b) was comparable to and the changes in four BA features (Figure 1j) was more dramatic than that of APOE-4 positive proportion during AD development. Four BA features in men were sensitive to the rate of MCI progression (Figures 3a-3e). These findings may offer novel insights into the roles that BAs might have at different stages in the development of AD and provide evidence for stage-specific diagnosis and treatment of AD.

Various types of BA features were selected as potential markers including the concentration of individual BA (CA), the proportion of individual BA to TBA (CA%), the ratios reflective of enzymatic activities and gut microbiome function (DCA/CA and GDCA/CA). Their early and continuous alterations indicate mechanistic roles of individual BA, BA pool composition, BA synthetic enzymes, and BA related gut microbiota in AD development. Of them, CA% is a newly identified one whereas the other three were identified previously by us [12, 13]. We and others have reported that there is considerable individual difference in BA pool [18, 21]. CA% reflects the composition of BA pool and thus may serve as a systematic supplement and validation to CA.

Sex differences were observed in BA profiles during AD development. TBA levels were observed to be higher in men than in women (Figure S2), which agrees with previous reports [21, 22]. The four selected BA features changed earlier and more dramatically in men than in women (Figure 2). Stronger associations between BA features and AD markers were observed in men than in women. The differences in BA profiles and changes between the sexes may be mediated by sex hormones because BAs and sex hormones share the same precursor, cholesterol [23]. In agreement with our findings, BA profiles have been shown to fluctuate with the initiation and progression of sexually dimorphic diseases, such as prostate cancer[24], ovarian cancer[25, 26], and vascular dementia[15]. Meanwhile, the sexual difference may due to different expressions of BA synthetic enzymes and transporters, and gut microbiota composition[23, 27, 28]. It is reported in animal studies that mice treated with estrogen or androgen have been shown to exhibit significant alterations in BA profiles [29]. Importantly, BAs have been used for the treatment of sex-specific diseases [24, 30, 31]. We previously reported the protective effect of bile acid sequestrants against vascular dementia in men but not in women [15]. Further studies are required to elucidate the mechanisms underlying sex differences in BA profiles and their effects on AD, which may pave the way for individualized medicine.

Previous research has suggested that disruptions in cholesterol homeostasis may increase the risk of developing AD [32-34]. We have proposed that changes in both serum and brain BA profiles may be involved in the development of AD and could be potential targets for the prevention and treatment of AD [16]. The results of this study support this hypothesis. The primary catabolic fate of brain and liver cholesterol is the conversion to primary BAs, CA and CDCA, through intermediate oxysterols (e.g. 24S-hydroxylcholesterol and 27-hydroxycholesterol in the brain and 25-hydroxycholesterol and 27-hydroxycholesterol in the liver) that can cross the blood-brain barrier [14, 16]. Our data showed that serum levels of these two primary BAs decreased gradually during the development of AD in men (Figure 2 and S3) and were associated with AD markers from CSF and brain imaging. We also found that primary BAs were detectable in the brain and observed downward trends in CA, CA%, and CDCA in the brains of male NC, MCI, and AD individuals from the ROSMAP cohort (Figure S4). In addition, we previously observed that patients taking BA sequestrants (drugs that reduce circulating BAs and increase cholesterol catabolism) had a lower incidence of all-cause dementia compared to patients taking other lipid-modifying therapies, using data from the UK Clinical Practice Research Datalink (CPRD) database [15]. Taken together, these findings suggest that the early declines in serum and brain levels of CA and CDCA may be due to abnormalities in cholesterol catabolism, and impaired primary BA synthesis may be an important factor in AD pathology.

The biosynthesis of BAs from cholesterol occurs via two major pathways, namely, the classic (neutral) pathway and the alternative (acidic) pathway[35]. The four BA features being identified as representative markers belong to the classical pathway. The levels of some BA features related to the alternative pathway (GLCA/CDCA, GLCA/UDCA, LCA/CDCA, LCA, GLCA, and GLCA%) were significantly altered in AD but not in EMCI and LMCI compared with NC (Table S10). Accordingly, abnormalities in the classical pathway may precede changes in the alternative pathway during AD progression. Unfortunately, we don’t have strong evidence to support this hypothesis and further study is needed to elucidate the dynamic balance of classic and alternative pathways in AD development.

Noninvasive and cost-effective markers and models are desirable for AD management. Blood marker-based models may be an attractive option. Our data demonstrate that the use of a simple logistic regression model comprising four BA features, three demographic markers, and APOE-4 status had comparable performance to models based on CSF and PET markers for the differentiation of NC and MCI (Figure 4c). Therefore, serum BA features may have utility in frequent and long-term monitoring to distinguish patients with MCI from high-risk NC individuals. We noticed that although the addition of BA features increased the performances of CSF and PET markers, the improvements were not as large as those of APOE-4 and demographics. This may be attributable to the small sample size of CSF and PET tests, with few individuals accepting PET imaging and CSF testing prior to progression to AD. Furthermore, the diagnostic and predictive powers of the CSF and PET panels were higher than that of BA features and increased with disease severity, thereby reducing the potential for improvement. In summary, our noninvasive model holds potential for clinical application, and the addition of BA markers may increase the performance of CSF and PET panels for MCI/AD diagnosis and MCI progression prediction.

This study has several limitations. Firstly, the identified BA markers and constructed models were specific to men, and it would be useful to investigate the same relationships in women or in both sexes. Secondly, we were unable to measure brain cholesterol levels, which are known to be associated with BA profiles and AD pathology. Thirdly, we used samples from multiple visits by the same subjects to increase statistical power, which may introduce bias. Additionally, the clinical potential of our findings may be limited due to the small number of individuals with CSF and PET markers. Despite these limitations, our preliminary observations on the changes in BA profiles using pooled samples and sex-stratified analyses offer novel insights that may help guide future pathological and clinical studies on AD. Therefore, it is important to validate our findings in larger, longitudinal studies with diverse ethnic and lifestyle patterns. It is also worth noting that other metabolites and metabolic pathways have been linked to AD development, and further research in these areas is warranted. Finally, our noninvasive model may be improved by adding other metabolic or serological markers.

In conclusion, our study highlights the significant sex differences and dynamic changes in BA profiles during AD initiation and progression. The inclusion of four BA features may improve the diagnostic and predictive capabilities of clinical marker panels for MCI and AD in men. Further research is needed to fully understand the gut microbiome-BA-brain cholesterol axis and identify potential targets for the prevention and treatment of AD.

## Data Availability

Metabolomics datasets used in the current analyses for the ADNI 1 and ADNI GO 2 cohorts are available via the Accelerating Medicines Partnership Alzheimer's Disease (AMP AD) Knowledge Portal and can be accessed at http://dx.doi.org/10.7303/syn5592519 (ADNI 1) and http://dx.doi.org/10.7303/syn9705278 (ADNI GO 2). The full complement of clinical and demographic data for the ADNI cohorts are hosted on the LONI data sharing platform and can be requested at http://adni.loni.usc.edu/data-samples/access-data/.

^1^

NC: Normal cognition
MCI: Mild cognitive impairment
EMCI: Early mild cognitive impairment
LMCI: Late mild cognitive impairment
AD: Alzheimer’s disease
sMCI: sustained MCI
cMCI: converted MCI
ADNI: Alzheimer’s Disease Neuroimaging Initiative
ROSMAP: Religious Orders Study and Rush Memory and Aging
BA: Bile acid
TBA: Concentration of total bile acids
CA: Cholic acid
CA%: Concentration percentage of cholic acid to total bile acids
DCA/CA: Ratio of deoxycholic acid and cholic acid
GDCA/CA: Ratio of deoxycholic acid glycine conjugate and cholic acid
CDCA: Chenodeoxycholic acid
A/T/N: Amyloid-beta, tau, and neurodegenerative
APOE-4: apolipoprotein E ε4 genotype
UK CPRD: Clinical Practice Research Datalink database of United Kingdom
AUC: Area under Receiver Operating Characteristic Curve
UPLC-MS/MS: ultra-performance liquid chromatography coupled to tandem mass spectrometry
PCA: Principle component analysis
LOESS: locally weighted regression

## Author Contributions

W.J. and R.K.-D. contributed to concept and design and led the team that included all co-authors. Drafting of the manuscript was done by T.C., B.S.C., W.J., and R.K.-D. T.C., B.S.C., M.J.S., T.S, M.A., S.M.D., K.N., and M.L. contributed to statistical analyses. L.W., G.X., T.C., K.B., and L.R. contributed to sample analysis and quality control. Data management and medication term mapping were done by M.A. Critical revision of the manuscript for important intellectual content was done by W.J., R.K.-D., B.S.C., M.A., M.J.S., Q.G., X.Z., A.K.P., and Z.R..

## Obtained funding

This work was supported by National Key R&D Program of China (2021YFA1301300) and National Natural Science Foundation of China (82270917, 81974073, and 31972935). Funding for ADMC (Alzheimer’s Disease Metabolomics Consortium, led by Dr R.K.D. at Duke University) was provided by the National Institute on Aging grant R01AG046171, a component of the Accelerating Medicines Partnership for AD (AMP AD) Target Discovery and Preclinical Validation Project (https://www.nia.nih.gov/research/dn/amp-ad-target-discovery-and-preclinical-validation-project) and the National Institute on Aging grant RF1 AG0151550, a component of the M2OVE-AD Consortium (Molecular Mechanisms of the Vascular Etiology of AD Consortium https://www.nia.nih.gov/news/decoding-molecular-ties-between-vascular-disease-and-alzheimers). Data collection and sharing for this project was funded by the Alzheimer’s Disease Neuroimaging Initiative (A.D.N.I.) (National Institutes of Health Grant U01 AG024904) and DOD A.D.N.I. (Department of Defense award number W81XWH-12-2-0012). A.D.N.I. is funded by the National Institute on Aging, the National Institute of Biomedical Imaging and Bioengineering, and through generous contributions from the following: AbbVie, Alzheimer’s Association; Alzheimer’s Drug Discovery Foundation; Araclon Biotech; BioClinica, Inc.; Biogen; Bristol-Myers Squibb Company; CereSpir, Inc.; Eisai Inc.; Elan Pharmaceuticals, Inc.; Eli Lilly and Company; EuroImmun; F. Hoffmann-La Roche Ltd and its affiliated company Genentech, Inc.; Fujirebio; GE Healthcare; IXICO Ltd.; Janssen Alzheimer Immunotherapy Research & Development, LLC.; Johnson & Johnson Pharmaceutical Research & Development LLC.; Lumosity; Lundbeck; Merck & Co., Inc.; Meso Scale Diagnostics, LLC.; NeuroRx Research; Neurotrack Technologies; Novartis Pharmaceuticals Corporation; Pfizer Inc.; Piramal Imaging; Servier; Takeda Pharmaceutical Company; and Transition Therapeutics. The Canadian Institutes of Health Research is providing funds to support A.D.N.I. clinical sites in Canada. Private sector contributions are facilitated by the Foundation for the National Institutes of Health (www.fnih.org). The grantee organization is the Northern California Institute for Research and Education, and the study is coordinated by the Alzheimer’s Disease Cooperative Study at the University of California, San Diego.

A.D.N.I. data are disseminated by the Laboratory for Neuro Imaging at the University of Southern California. A complete listing of A.D.N.I. investigators who contributed to the design and implementation and/or provided data but did not participate in analysis or writing of this report can be found at: http://adni.loni.usc.edu/wp-content/uploads/how_to_apply/ADNI_Acknowledgement_List.pdf.

## Harmonization of methods

Alzheimer’s Disease Metabolomics Consortium (ADMC): A complete listing of ADMC investigators can be found at: https://sites.duke.edu/adnimetab/who-we-are/.

## Role of the funder/sponsor

Funders had no role in the design and conduct of the study; collection, management, analysis, and interpretation of the data; preparation, review, or approval of the manuscript; and decision to submit the manuscript for publication.

## Additional Contributions

The authors are grateful to Lisa Howerton for administrative support and the numerous ADNI study volunteers and their families.

## Conflicts

The authors declare no conflict of interest.

## Supplementary information

1. Study cohorts and sample collection
2. Quantitative measurement of metabolites
3. Quality control and pretreatment of metabolic data
4. Clinical marker panels
5. Statistical analyses
6. Brain BA levels of ROS/MAP cohort
7. Abbreviation
8. SI Figures Figure S1. Dynamic changes (LOESS fitted curves with 95% CI of PC1, the first components of PCA) of a) 104 serum metabolites and b) different types of metabolites during AD progression. Metabolites start to change early in AD development and continue across AD stages. Changes of BAs are more apparent than that of other types of metabolites. Figure S2. TBA levels (mean with S.E.) in NC and all samples (a) and in the four diagnostic groups (b) stratified by sex. *, significant difference based on the Kruskal– Wallis test with post-hoc comparisons. Figure S3. Levels (mean with S.E.) of CDCA (a) and CDCA% (b) in the four diagnostic groups stratified by sex. *, significant difference based on the Kruskal–Wallis test with post-hoc comparisons. Figure S4. Levels (mean with S.E.) of CA (a), CA% (b), and CDCA (c) in NC, MCI, and AD stratified by sex in brain samples from the ROS/MAP cohort. There is no statistical significance among or between groups, presumably due to the small sample size.
9. SI tables Table S1. BA features evaluated in this study. Table S2. Medications selected for adjustment for BA features. Table S3. Demographic and clinical characteristics of samples stratified by visit. Table S4. Demographic and clinical characteristics of samples stratified by MCI progression to AD. Table S5. Association of BA features with MCI progression to AD in men and women (P-values and OR with 95% CI from logistic regression). Table S6. Demographic and clinical characteristics of samples with progression stratified by months before MCI/AD and months after AD. Table S7. Performances of logistic regression models in discriminating NC, MCI, and AD based on clinical marker panels alone and combined with four BA features in men. Table S8. Performances of MCI to AD prediction logistic regression models based on clinical marker panels alone and combined with four BA features in men. Table S9. Performances of diagnostic and predictive models using eight noninvasive markers. Table S10. Levels (mean with S.E.) of features related to alternative pathway in four diagnostic groups.

## Notes

### Competing Interest Statement

The authors have declared no competing interest.

## Reference

[1] Dumitrescu L, Barnes LL, Thambisetty M, Beecham G, Kunkle B, Bush WS, et al. Sex differences in the genetic predictors of Alzheimer’s pathology. Brain. 2019;142:2581–9.

[2] Yan S, Zheng C, Paranjpe MD, Li Y, Li W, Wang X, et al. Sex modifies APOE epsilon4 dose effect on brain tau deposition in cognitively impaired individuals. Brain. 2021;144:3201–11.

[3] Lirong W, Mingliang Z, Mengci L, Qihao G, Zhenxing R, Xiaojiao Z, et al. The clinical and mechanistic roles of bile acids in depression, Alzheimer’s disease, and stroke. Proteomics. 2022;22:e2100324.

[4] Wang J, Wei R, Xie G, Arnold M, Kueider-Paisley A, Louie G, et al. Peripheral serum metabolomic profiles inform central cognitive impairment. Scientific reports. 2020;10:14059.

[5] Cui M, Jiang Y, Zhao Q, Zhu Z, Liang X, Zhang K, et al. Metabolomics and incident dementia in older Chinese adults: The Shanghai Aging Study. Alzheimer’s & dementia : the journal of the Alzheimer’s Association. 2020;16:779–88.

[6] Kim M, Snowden S, Suvitaival T, Ali A, Merkler DJ, Ahmad T, et al. Primary fatty amides in plasma associated with brain amyloid burden, hippocampal volume, and memory in the European Medical Information Framework for Alzheimer’s Disease biomarker discovery cohort. Alzheimer’s & dementia : the journal of the Alzheimer’s Association. 2019;15:817–27.

[7] Toledo JB, Arnold M, Kastenmüller G, Chang R, Baillie RA, Han X, et al. Metabolic network failures in Alzheimer’s disease: A biochemical road map. Alzheimer’s & dementia : the journal of the Alzheimer’s Association. 2017;13:965–84.

[8] Arnold M, Nho K, Kueider-Paisley A, Massaro T, Huynh K, Brauner B, et al. Sex and APOE ε4 genotype modify the Alzheimer’s disease serum metabolome. Nature communications. 2020;11:1148.

[9] Zheng X, Chen T, Zhao A, Ning Z, Kuang J, Wang S, et al. Hyocholic acid species as novel biomarkers for metabolic disorders. Nature communications. 2021;12:1487.

[10] Zheng X, Chen T, Jiang R, Zhao A, Wu Q, Kuang J, et al. Hyocholic acid species improve glucose homeostasis through a distinct TGR5 and FXR signaling mechanism. Cell Metab. 2021;33:791–803 e7.

[11] Nie K, Li Y, Zhang J, Gao Y, Qiu Y, Gan R, et al. Distinct Bile Acid Signature in Parkinson’s Disease With Mild Cognitive Impairment. Front Neurol. 2022;13:897867.

[12] MahmoudianDehkordi S, Arnold M, Nho K, Ahmad S, Jia W, Xie G, et al. Altered bile acid profile associates with cognitive impairment in Alzheimer’s disease-An emerging role for gut microbiome. Alzheimer’s & dementia : the journal of the Alzheimer’s Association. 2019;15:76–92.

[13] Nho K, Kueider-Paisley A, MahmoudianDehkordi S, Arnold M, Risacher SL, Louie G, et al. Altered bile acid profile in mild cognitive impairment and Alzheimer’s disease: Relationship to neuroimaging and CSF biomarkers. Alzheimer’s & dementia : the journal of the Alzheimer’s Association. 2019;15:232–44.

[14] Baloni P, Funk CC, Yan J, Yurkovich JT, Kueider-Paisley A, Nho K, et al. Metabolic Network Analysis Reveals Altered Bile Acid Synthesis and Metabolism in Alzheimer’s Disease. Cell Rep Med. 2020;1:100138.

[15] Varma VR, Wang Y, An Y, Varma S, Bilgel M, Doshi J, et al. Bile acid synthesis, modulation, and dementia: A metabolomic, transcriptomic, and pharmacoepidemiologic study. PLoS medicine. 2021;18:e1003615.

[16] Jia W, Rajani C, Kaddurah-Daouk R, Li H. Expert insights: The potential role of the gut microbiome-bile acid-brain axis in the development and progression of Alzheimer’s disease and hepatic encephalopathy. Medicinal research reviews. 2020;40:1496–507.

[17] Chang R, Trushina E, Zhu K, Zaidi SSA, Lau BM, Kueider-Paisley A, et al. Predictive metabolic networks reveal sex- and APOE genotype-specific metabolic signatures and drivers for precision medicine in Alzheimer’s disease. Alzheimer’s & dementia : the journal of the Alzheimer’s Association. 2022.

[18] Xiang X, Backman JT, Neuvonen PJ, Niemi M. Gender, but not CYP7A1 or SLCO1B1 polymorphism, affects the fasting plasma concentrations of bile acids in human beings. Basic & clinical pharmacology & toxicology. 2012;110:245–52.

[19] Xie G, Wang L, Chen T, Zhou K, Zhang Z, Li J, et al. A Metabolite Array Technology for Precision Medicine. Anal Chem. 2021;93:5709–17.

[20] Petersen RC. Alzheimer’s disease: progress in prediction. Lancet Neurol. 2010;9:4–5.

[21] Xie G, Wang Y, Wang X, Zhao A, Chen T, Ni Y, et al. Profiling of serum bile acids in a healthy Chinese population using UPLC-MS/MS. J Proteome Res. 2015;14:850–9.

[22] Montagnana M, Danese E, Giontella A, Bonafini S, Benati M, Tagetti A, et al. Circulating Bile Acids Profiles in Obese Children and Adolescents: A Possible Role of Sex, Puberty and Liver Steatosis. Diagnostics (Basel). 2020;10.

[23] Phelps T, Snyder E, Rodriguez E, Child H, Harvey P. The influence of biological sex and sex hormones on bile acid synthesis and cholesterol homeostasis. Biol Sex Differ. 2019;10:52.

[24] Gafar AA, Draz HM, Goldberg AA, Bashandy MA, Bakry S, Khalifa MA, et al. Lithocholic acid induces endoplasmic reticulum stress, autophagy and mitochondrial dysfunction in human prostate cancer cells. PeerJ. 2016;4:e2445.

[25] Jin Q, Noel O, Nguyen M, Sam L, Gerhard GS. Bile acids upregulate BRCA1 and downregulate estrogen receptor 1 gene expression in ovarian cancer cells. Eur J Cancer Prev. 2018;27:553–6.

[26] Horowitz NS, Hua J, Powell MA, Gibb RK, Mutch DG, Herzog TJ. Novel cytotoxic agents from an unexpected source: bile acids and ovarian tumor apoptosis. Gynecol Oncol. 2007;107:344–9.

[27] Markle JG, Frank DN, Mortin-Toth S, Robertson CE, Feazel LM, Rolle-Kampczyk U, et al. Sex differences in the gut microbiome drive hormone-dependent regulation of autoimmunity. Science. 2013;339:1084–8.

[28] Org E, Mehrabian M, Parks BW, Shipkova P, Liu X, Drake TA, et al. Sex differences and hormonal effects on gut microbiota composition in mice. Gut Microbes. 2016;7:313–22.

[29] Zhang B, Shen S, Gu T, Hong T, Liu J, Sun J, et al. Increased circulating conjugated primary bile acids are associated with hyperandrogenism in women with polycystic ovary syndrome. J Steroid Biochem Mol Biol. 2019;189:171–5.

[30] Schuldes H, Dolderer JH, Zimmer G, Knobloch J, Bickeboller R, Jonas D, et al. Reversal of multidrug resistance and increase in plasma membrane fluidity in CHO cells with R-verapamil and bile salts. Eur J Cancer. 2001;37:660–7.

[31] Lee WS, Jung JH, Panchanathan R, Yun JW, Kim DH, Kim HJ, et al. Ursodeoxycholic Acid Induces Death Receptor-mediated Apoptosis in Prostate Cancer Cells. J Cancer Prev. 2017;22:16–21.

[32] Xue-Shan Z, Juan P, Qi W, Zhong R, Li-Hong P, Zhi-Han T, et al. Imbalanced cholesterol metabolism in Alzheimer’s disease. Clin Chim Acta. 2016;456:107–14.

[33] van der Kant R, Langness VF, Herrera CM, Williams DA, Fong LK, Leestemaker Y, et al. Cholesterol Metabolism Is a Druggable Axis that Independently Regulates Tau and Amyloid-beta in iPSC-Derived Alzheimer’s Disease Neurons. Cell Stem Cell. 2019;24:363–75 e9.

[34] Polidori MC, Pientka L, Mecocci P. A review of the major vascular risk factors related to Alzheimer’s disease. J Alzheimers Dis. 2012;32:521–30.

[35] Jia W, Wei M, Rajani C, Zheng X. Targeting the alternative bile acid synthetic pathway for metabolic diseases. Protein Cell. 2021;12:411–25.

